# Robust immediate and limited long-term benefit of prism adaptation on spatial neglect: A Systematic Review and Meta-Analysis of outcomes and predictors

**DOI:** 10.1101/2025.06.09.25329277

**Authors:** YuanLiang Zhu, Francois Quesque, Daisuke Nishida, Sophie Jacquin-Courtois, Jacques. Luaute, Eric Chabanat, Gilles Rode, Yves Rossetti

## Abstract

**Objective:** The first aim was to assess the immediate and long-term effects of prism adaptation (PA) on spatial neglect (SN) via meta-analysis of randomized controlled trials (RCTs). The second aim was to identify potential predictive factors of PA efficacy.

**Methods:** We searched 4 databases up to March 2025 for RCTs comparing PA or PA+conventional vs. shamPA or shamPA+conventional or only conventional in participants with SN, and assessing behavioral or neuropsychological tests (such as Catherine Bergego Scale (CBS) and Behavioral Inattention Test (BIT)). Cochrane risk-of-bias assessment tool and random-effects meta-analysis were used, and effect size was reported as Weighted Mean Difference (WMD) with 95%CI.

**Results:** Only 10 RCTs (356 participants) could be included, 8 of them reporting CBS data and 6 for BIT. Immediate PA effects on SN showed a significant improvement for CBS between the PA group and control group (all studies with prism shift ≥10°, WMD= −2.13, 95%CI: [−3.93, −0.33], P<0.05), while non-significance for BIT-C. Long-term benefit was not significant for CBS, while significant for BIT-C when the prism shift was ≥10° (WMD= 12.37, 95%CI: [0.53, 24.21], P<0.05). Linear regressions showed non-significant predictive factors among the participant characteristics or PA intervention parameters. Subgroup analyses for CBS showed a significant immediate improvement in the larger total quantity of prism exposure (number of total trials × prism shift) subgroup (WMD= −2.73, 95%CI: [−5.01, −0.44], P<0.05), whereas subgroup with smaller total exposure showed non-significance.

**Conclusion:** Even with stringent inclusion criteria, robust effects of PA on SN were observed in the short-term CBS (all studies with prism shift ≥10°), mainly derived from studies with total exposure ≥11250°·trials. A significant improvement for long-term BIT-C was observed when the prism shift was ≥10° (n=2, with 4000°·trials and 21600°·trials). The total quantity of prism exposure (°·trials) may be a useful predictive efficacy factor of efficacy.

## 1. Introduction

Spatial neglect (SN), a lateralized neuropsychological disorder of spatial cognition, has a prevalence of over 50% in the first weeks and months after a stroke[1]. It may result in an abnormal bias toward the space ipsilateral to the injured cerebral hemisphere, so that people pay little or no attention to the contralateral side (not a primary sensory or motor deficit) to real or imagined space, and are less likely to orient sensory receptors and actions toward this side of space[2, 3]. SN (rather than overall stroke severity) predicts poor functional recovery, prolonged hospitalization, functional dependence, disability in life activities, and increased risk of falls[4–6]. With rising post-acute care for stroke survivors, the need for validated SN rehabilitation is growing[7].

Prism adaptation (PA), a promising intervention to reduce SN symptoms and improve functional prognosis, was first applied to SN rehabilitation by Rossetti and colleagues in 1998[8–12]. They found that the signs of SN on visuomotor tasks significantly improved in the PA group after a few minutes of prism exposure to a right-shifted prism, compared to the control group[8]. In the early phase (exposure) of the classic PA paradigm, individuals’ entire visual field will be deflected due to the refraction of prisms (optical shift), and their reaching movements are biased toward the virtual target when they are asked to point at a visual target (direct effect of prisms). Over trial repetition, they can gradually improve performance through error reduction until restoring a normal level of performance (error compensation process). When prisms are removed, visuomotor errors in the direction opposite to the optical shift should be observed (after-effect) and measured to quantify the amount of adaptation[12]. This intervention is thought to bias the egocentric reference and increase spatial perception/attention on the affected side of participants with SN[9]. In recent years, some studies on the underlying neurophysiological mechanisms of PA have gradually revealed how it works[13]. These studies suggest that PA is not only a perceptual-motor corrective process, but also involves complex neural network remodeling, including the restoration of interhemispheric functional balance, attention and sensorimotor network remodeling[13–15]. Indeed, generalization of PA after-effects has been reported for numerous domains such as wheel-chair driving[16], dichotic listening[17], and beyond SN to posture[18], spatial dysgraphia[19], or number cognition[20]. This extraordinary expansion from a sensory-motor process to cognitive representations allowed the prediction of a good generalization of PA to neglect rehabilitation[21]. Accordingly, improvement of activities of daily living (ADL) has also been reported e.g.[22, 23]. However, case reports and randomized controlled trials (RCTs, widely regarded as the gold standard of study design in health and medical research) do not necessarily lead to similar conclusions[24].

In the past 27 years since PA was first applied in SN, a lot of RCTs have been conducted[25–28]. However, the results of these studies remain controversial and insufficiently consistent[29]. Therefore, it is of prime importance to determine the short-term beneficial effects and long-term efficacy of PA on SN[30]. Three meta-analyses related to this topic have been published previously, with data extracted up to 30 December 2019, 31 January 2020 and 31 June 2021, respectively[29, 31, 32]. Several new high-quality RCTs with inconsistent results have been published in recent years[27, 28, 33, 34]. Given the current debate on PA efficacy on SN and the stake of its concrete clinical implications, it is necessary and timely to re-examine and comprehensively review the available evidence based on the most rigorous and reliable source of information, i.e. randomized controlled clinical trials. During the revision process of the present report, a related meta-analysis published in March 2025 (data extracted up to April 2024), reported a significant short-term effect of PA intervention on SN outcomes (BIT-C and cancellation tasks), and found that larger prism shift (≥10° shift) was a crucial factor in eliciting PA intervention effects[35]. This has important guiding significance for future clinical research. However, this meta-analysis mainly focused on the short-term effect of PA on SN, and the impact of intervention timing (acute or chronic stroke), PA protocol (concurrent PA or terminal PA), and a single PA intervention parameter (prism magnitude) on the PA efficacy on SN.

Therefore, we aimed to make an important expansion based on previous meta-analyses. We performed a meta-analysis specifically including RCTs published before 2025 and distinguished between immediate and long-term effects of PA on SN. Two main sources of variability of PA efficacy have been identified: the physical parameters of PA intervention (e.g., prism strength, number of sessions etc.), and the participant characteristics (especially the variety of brain lesion topography and size). Therefore, our secondary goal was to identify which of the physical parameters of PA interventions provide a predictive value of its efficacy. Notably, PA efficacy may not only be influenced by a single parameter but may also be related to the overall intervention dose (e.g., ‘total quantity of prism exposure’)[36]. Additionally, the impact of available participant characteristics on the PA efficacy on SN was analyzed to search for further guidance for future studies and clinical applications.

## 2. Methods

The authors followed the PRISMA2020 and Cochrane guidelines for Systematic Reviews of Interventions. This meta-analysis study was registered in Prospero in February 2025 (CRD420250650402).

### 2.1 Literature search

Based on PRISMA guidelines, a comprehensive literature search was performed in four databases (PubMed, Cochrane, Embase, and Web of Science) up to March 2025[37]. The search terms were (‘prism adaptation’ OR ‘prism exposure’ OR ‘prism glasses’) AND (‘unilateral neglect OR ‘unilateral spatial neglect’ OR ‘hemispatial neglect’ OR ‘visuospatial neglect’ OR ‘neglect’). The search was conducted by two authors (YLZ, FQ) independently, and no disparity was identified.

### 2.2 Selection criteria and study selection

Two authors (YLZ, FQ) independently screened the studies based on the inclusion criteria. A third author (YR) solved potential disagreements. The inclusion criteria were as follows according to PICOS: P (participants): participants with SN; I (intervention): PA/PA + conventional treatment; C (comparison): sham PA/sham PA + conventional treatment/only conventional treatment; O (outcome): containing behavioral or neuropsychological tests, such as the Catherine Bergego Scale (CBS), Behavioral Inattention Test (BIT), Functional Independence Measure (FIM), Dynamic and Static Task; S (study design): RCT. The exclusion criteria were as follows: no PA; no baseline measurement; no RCT; no behavioral or neuropsychological assessment; data not available or without mean, SD/SEM; Excluded studies included review article, meta-analysis, case report, protocol, meeting, summary, conference abstract, and animal trials.

### 2.3 Data extraction

Two authors (YLZ, YR) independently extracted the following data: study design, the number of participants, age, gender, time post-stroke/injury, intervention in each group, PA intervention parameters, and outcomes (assessment time).

### 2.4 Outcome assessment

During article screening, we did not intentionally restrict the type of outcomes and included all studies that reported behavioral or neuropsychological tests, such as CBS, BIT, Bells Test, scene copying test, FIM, Dynamic Task (Wheelchair Navigation Task), and Static Task (shape cancellation task). As the number of data points available for other reported outcomes was insufficient to support meaningful meta-analysis (n ≤ 2, Table S1), we only focused on CBS and BIT, which are also the two most commonly used scales for assessing neglect. The CBS could assess aspects of participants’ perceptual and motor dysfunction separately, as well as awareness of the impact of SN on daily living skills[38, 39]. It has been shown to have good reliability, validity and sensitivity to changes in the rehabilitation process of participants with SN[40]. Therefore, it was chosen as the main outcome in our study. The BIT was widely used to assess the perceptual and spatial cognitive motor systems[41]. Considering its insufficient sensitivity and limited ecological validity, it was used as the secondary outcome in our study[41, 42].

#### 2.4.1 Main outcome

The CBS includes 10 real-life situations that capture various aspects of ADL (grooming, eating, dressing etc.), with each task ranging from 0 points (no neglect) to 3 points (severe neglect), and with a final maximum score of 30 points[38, 39].

#### 2.4.2 Secondary outcome

The BIT is usually divided into two categories: conventional BIT (BIT-C) and behavioral BIT (BIT-B)[43]. BIT-C is used to assess the severity of visual neglect and has 6 items with a maximum score of 146; while BIT-B assesses ADLs and has 9 items with a maximum score of 81. The lower the score, the more severe the visual impairment, and a total BIT score of less than 196 indicates SN.

### 2.5 Assessment of risk of bias

The Cochrane risk-of-bias assessment tool was used to assess potential risk bias[44]. Two authors (YLZ, DN) independently assessed the risk of bias. A third author (EC) joined in the discussion to resolve disagreements, achieving consensus in all cases.

### 2.6 Meta-analysis

Meta-analysis was performed using Stata 16.0, based on the evaluation time after intervention: immediate (<24 hours after the end of treatment) and long-term (>1 month after the end of treatment). Data were averaged when multiple data points were available at a certain time.

Treatment effects were reported as Weighted Mean Differences (WMD) with 95% confidence intervals (CI)[45]. The I^2^ statistic was used to assess heterogeneity, with 0% indicating no observed heterogeneity, >25%, >50%, and >75% indicating low, moderate, and high heterogeneity, respectively[46]. Considering the small sample size, and clinical heterogeneity across studies (e.g. participant characteristic, study design, or different PA intervention parameters), we employed a random-effects model for all meta-analyses independent of observed I² statistics[45].

### 2.7 Sensitivity analyses

In Stata 16.0, we used influence analysis and leave-one-out analysis to execute sensitivity analyses, to assess the robustness and reliability of the combined results, and to further discuss potential sources of heterogeneity and their impact on the results.

### 2.8 Publication bias

Publication bias could be assessed by using funnel plot asymmetry testing (including Egger’s test) when ≥10 studies were included[45], while this threshold was not met in our study (n ≤7 in short-/long-term).

### 2.9 Univariate linear regression analyses

Due to the limited number of BIT-C reports (only three studies each included in short- and long-term), we only conducted univariate linear regression analyses (JASP-Jeffreys’s Amazing Statistics Program software) for the main outcome-CBS, with the largest number of studies included. To exploratorily identify whether participants’ basic characteristics (average age and days after stroke) and specific PA intervention parameters, i.e. prism shift (°), number of trials (during prism exposure), number of sessions of each study, total number of trials (number of trials × number of sessions), frequency (/day), number of PA intervention days, total duration of the PA intervention (days elapsed between the first-to-last sessions), and the total quantity of prism exposure (hereafter referred to as “total exposure”, calculated as number of total trials × prism optical shift) might predict CBS improvement.

When available, individual results of short- and long-term CBS changes were calculated as follows: Result = ΔPA group – ΔControl group, i.e., ΔPA group = (Post-intervention value of PA group – Pre-intervention value of PA group), ΔControl group = (Post-intervention value of control group – Pre-intervention value of control group). As the total number of data points was only 11, the short-term and long-term CBS scores were pooled in this tentative analysis. Each study’s sample size was used as a weight variable in the analyses.

One study was excluded from the regression analyses for the number of trials, number of total trials and total exposure due to missing information on specific trial numbers per session[34]. Based on the available information (20 sessions of 20 min exposure to a 10° shift), we assumed that much more than 57 trials were achieved in 20 mins (e.g., 60 trials in less than 10 min in Rossetti et al. 1998), i.e. >11250°·trials. Thus, it was included in the larger total exposure group in the subgroup analyses.

### 2.10 Subgroup analyses

Although no significant predictive factors were identified by regression analyses (see Results 3.4 section), we conducted exploratory subgroup analyses for two PA intervention parameters based on prior evidence.

The first parameter was prism shift, based on the dose-response relationship identified in the most recent meta-analysis (Naito et al. 2025), suggesting a crucial role of larger prism shift (≥10°) in the effects of PA[35]. Studies were accordingly categorized into two subgroups: larger prism shift (≥10°) and smaller prism shift (<10°) subgroups. The second parameter was total exposure (number of total trials × prism shift), based on the idea that it could provide insight into the overall PA intervention dosage (Michel et al. 2007)[36]. According to half of its maximum value (22500°·trials) among the included studies, studies were divided into two subgroups: larger (≥11250°·trials) and smaller total exposure (<11250°·trials). Given the limited sample size for BIT (n ≤ 3 for each subgroup), we only performed regime subgroup analyses on CBS.

## 3. Results

### 3.1 Characteristics of the studies

In total, 1394 articles were found after literature searching, and 731 articles remained after excluding duplicate articles. Based on the inclusion and exclusion criteria, 14 articles were finally included. Original data from three articles remained unavailable after we contacted the corresponding author twice. In one of the articles, no information was provided about the randomization of participants. Therefore, we gathered comprehensive information from 10 remaining articles (published from 2008 to 2023)[25–28, 33, 34, 47–50]. The data screening process is shown in Fig. 1, and the basic characteristics of the included literature are shown in Table 1.

**Fig. 1.**
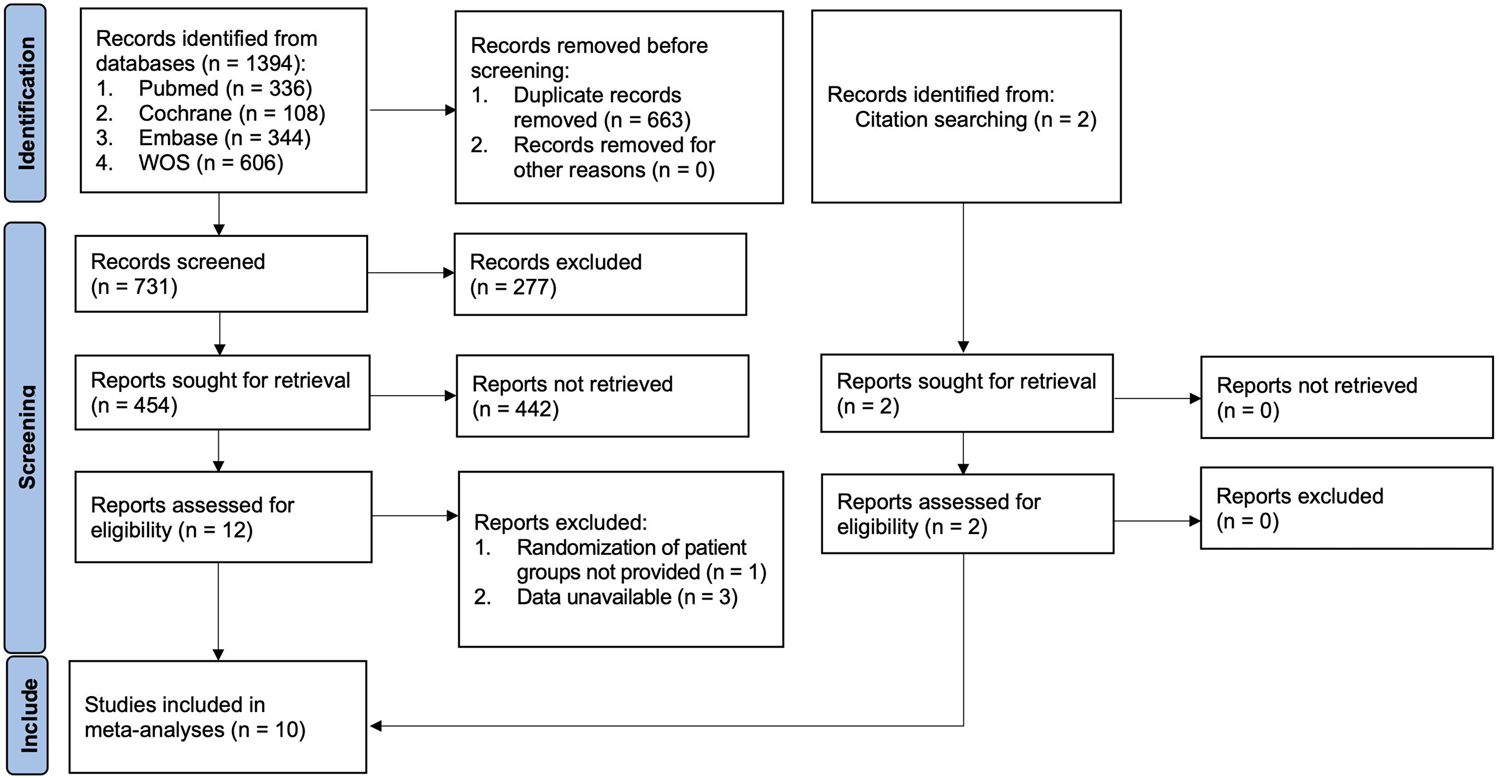
Flowchart of the study screening process. Out of the 1394 studies considered, only 10 were finally included in the present analysis.

**Table 1:** Basic characteristics of the included articles.

| Studies | Disease type<br>(hemisphere<br>side) | Groups |  |  |  |  |  |  |  |  |  | PA intervention parameters | Outcomes: CBS/BIT only<br>(Assessment time) |
| --- | --- | --- | --- | --- | --- | --- | --- | --- | --- | --- | --- | --- | --- |
|  |  | PA group |  |  |  |  | Control group |  |  |  |  |  |  |
|  |  | Sample<br>size | Age:<br>mean<br>(SD)/range | Gender<br>(M/F) | Mean time<br>post stroke<br>(days): mean<br>(SD)/range | Intervention | Sample<br>size | Age:<br>mean<br>(SD)/range | Gender<br>(M/F) | Mean time<br>post stroke<br>(days): mean<br>(SD)/range | Intervention |  |  |
| Nys 2008[25] | stroke- neglect<br>(R hemisphere) | 10 | 63.6 (13.8) | 7/3 | 8.8 (5.3) | 10°-right | 6 | 61.5 (11.0) | 3/3 | 11.2 (6.4) | 0° | 30mins/session,<br>4 consecutive days<br>Total sessions: 4<br>Total exposure<sup>#</sup>: 4000°·trials | Immediate: No<br>Long term: BIT-C<br>(1 month after the end of treatme |
| Turton 2010[47] | stroke- neglect<br>(R hemisphere) | 16 | 72 (14) | 8/8 | 45 (23) | 6°-right | 18 | 71 (14) | 11/7 | 47 (39) | 0° | Once/weekday, for 2 weeks<br>Total sessions: 10<br>Total exposure: 5400°·trials | Immediate: No<br>Long term: CBS, BIT-C<br>(8 weeks after the end of treatme |
| Mizuno 2011[48]<br>(multicenter, double<br>blind) | stroke- neglect<br>(R hemisphere) | 18 | 66.0 (11.5) | 12/6 | 67.1 (18.4) | 12°-right | 20 | 66.6 (7.7) | 15/5 | 64.4 (20.9) | 0° | 20 mins/session, twice/day,<br>5 days/week, for 2 weeks<br>Total sessions: 20<br>Total exposure: 21600°·trials | Immediate: CBS, BIT-C, BIT-B<br>(after the end of treatment)<br>Long term: CBS, BIT-C, BIT-B<br>(discharge) |
| Rode 2015[26]<br>(double blind) | stroke- neglect<br>(R hemisphere) | 9 | 55.2 (11.9) | 5/4 | 51 (17.4) | 10°-right | 9 | 61.7 (12.9) | 5/4 | 52.2 (19.3) | 0° | 6–10 mins/session, once/week,<br>for 4 weeks<br>Total sessions: 4<br>Total exposure: 3200°·trials | Immediate: No<br>Long term: total BIT<br>(5 months after the end<br>treatment) |
| Mancuso 2016[49]<br>(multicenter) | stroke- neglect<br>(R hemisphere) | 23 | 67.48 (9.40) | 17/6 | 66.3 (43.52) | 10°-right | 17 | 62.88 (12.55) | 11/6 | 51.24 (26.67) | 0° | 20 mins/session, twice/day,<br>5 days/week, for two weeks<br>Total sessions: 20<br>Total exposure: 18000°·trials | Immediate: CBS, BIT-C<br>(after the end of treatment)<br>Long term: No |
| Ten Brink 2017[50]<br>(double blind) | stroke- neglect<br>(R, L hemisphere,<br>bilateral) | 34 | 59.31 (14.45) | 25/9 | 41.5 (39.00) | 10°-right | 35 | 61.48 (13.37) | 24/11 | 37 (37) | 0° | Once/working day, for 2 weeks<br>Total sessions: 10<br>Total exposure: 10000°·trials | Immediate: CBS<br>(after the end of treatment)<br>Long term: CBS<br>(4 weeks after the end of treatme |
| Choi 2019[33] | stroke- neglect<br>(R hemisphere) | 10 | 62.90 (8.64) | 6/4 | 30 days:1<br>30-60 days: 6<br>60-90 days: 3 | 15°-right + FES | 10 | 66.00 (12.09) | 3/7 | 30 days:2<br>30-60 days: 5<br>60-90 days: 3 | only FES | 20 mins (15° PE + FES)/day,<br>5 times/week, for 3 weeks<br>Total sessions: 15<br>Total exposure: 22500°·trials | Immediate: CBS<br>(after the end of treatment)<br>Long term: No |
| Vilimovsky 2021[27]<br>(double blind) | 21 stroke-neglect<br>2 TBI-neglect<br>(R/L hemisphere) | 12 | 47.5-55 | 5/7 | 38.5-74 | 11.4°-right | 11 | 53–61 | 5/6 | 35–79 | 0° | 15–20 minutes/once,<br>10 sessions for 2 weeks<br>Total sessions: 10<br>Total exposure: 6840°·trials | Immediate: CBS<br>(after the end of treatment)<br>Long term: CBS<br>(4 weeks after the end of treatme |
| Choi 2022[34]<br>(multicenter) | stroke- neglect<br>(R hemisphere) | 12 | 62.9 (8.64) | 7/5 | 267.6 (79.2) | 10°-right + NV | 12 | 67.70 (9.76) | 4/8 | 291 (94.8) | only NV | 20 mins (10° PE + NV)/ day,<br>5 times/week, for 4 weeks<br>Total sessions: 20<br>Total exposure: 20000°·trials | Immediate: CBS<br>(after the end of treatment)<br>Long term: No |
| Umeonwuka 2023[28]<br>(double blind) | stroke- neglect<br>(R hemisphere) | 37 | 51.78 (13.06) | 17/20 | 26-56 | 11.4°-right | 37 | 46.51 (13.56) | 20/17 | 28–47 | 0° | 30 mins/session, once/day,<br>every weekday, over 16 days<br>Total sessions: 12<br>Total exposure: 12312°·trials | Immediate: CBS, BIT-C<br>(after the end of treatment)<br>Long term: No |

<sup>#</sup>: Total quantity of prism exposure (number of total trials x prism shift) BIT: Behavioral Inattention Test; BIT-B: behavioral BIT; BIT-C: conventional BIT; CBS: Catherine Bergego Scale; FES: functional electrical stimulation; L: left; NV: neck vibration; PA: prism adaptation; PE: prism exposure; R: right; TBI: traumatic brain injury.

### 3.2 Risk-of-bias assessment results

All 10 articles included in this study are RCTs on the effect of PA on SN, and five articles among them are double-blind[26, 28, 48, 50]. Six articles had incomplete data, but most of them explained the tenable reasons and reported sufficient data to calculate the standardized effect size for the outcome measures (Fig. 2).

**Fig. 2.**
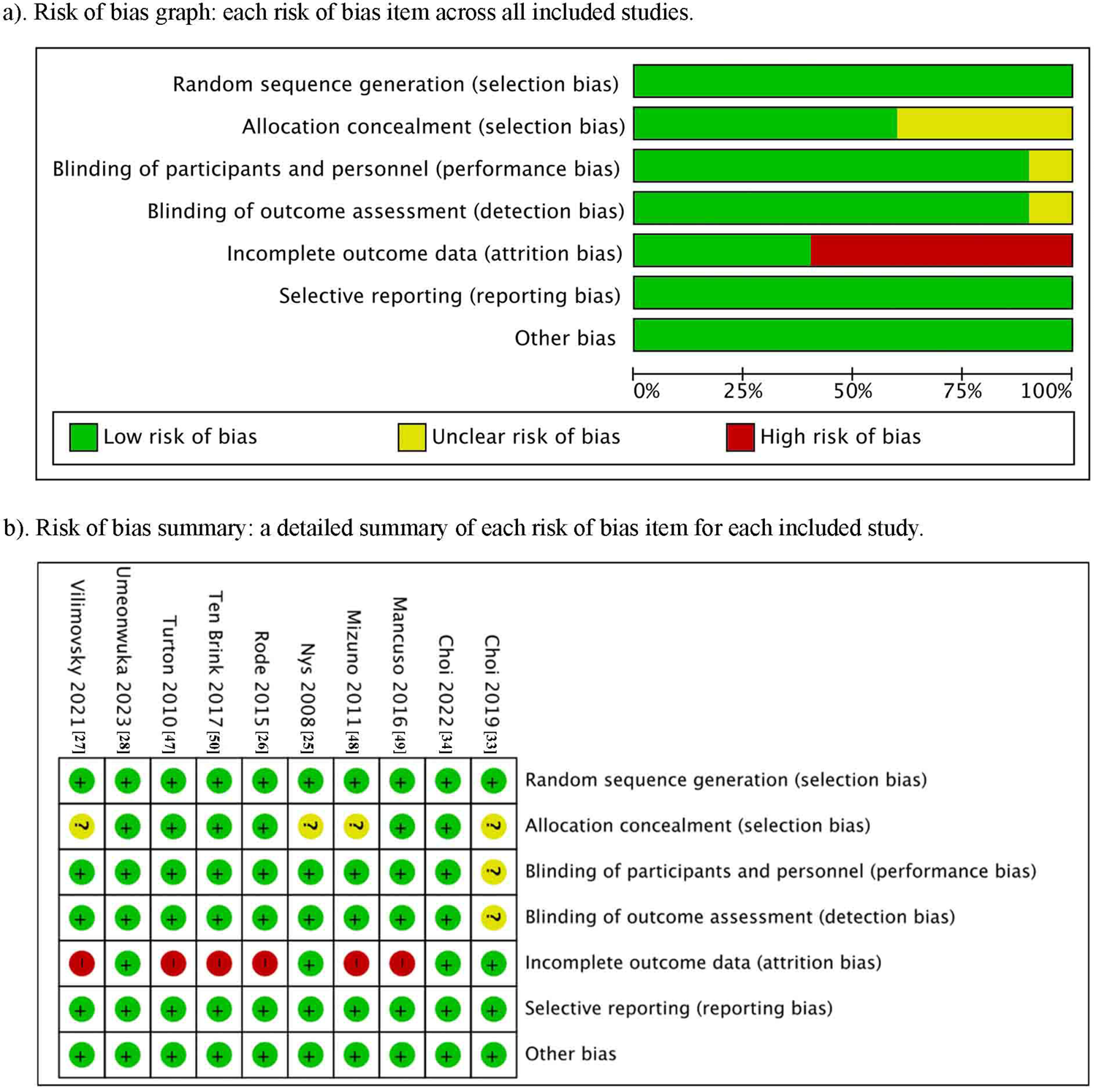
The risk-of-bias. a) Risk of bias graph: each risk of bias item across all included studies. b) Risk of bias summary: a detailed summary of each risk of bias item for each included study.

### 3.3 Outcomes

#### 3.3.1 Main outcome—CBS

For the immediate effect of PA intervention, a total of seven articles were included, with a total of 288 participants, of which 146 were in the PA group (average CBS score improvement: 6.53) and 142 were in the control group (average CBS score improvement: 4.08)[27, 28, 33, 34, 48–50]. As shown in Fig. 3a, the meta-analysis found that the PA group showed greater improvement than the sham group (WMD= −2.13, 95% CI: [−3.93, −0.33], P <0.05).

**Fig. 3.**
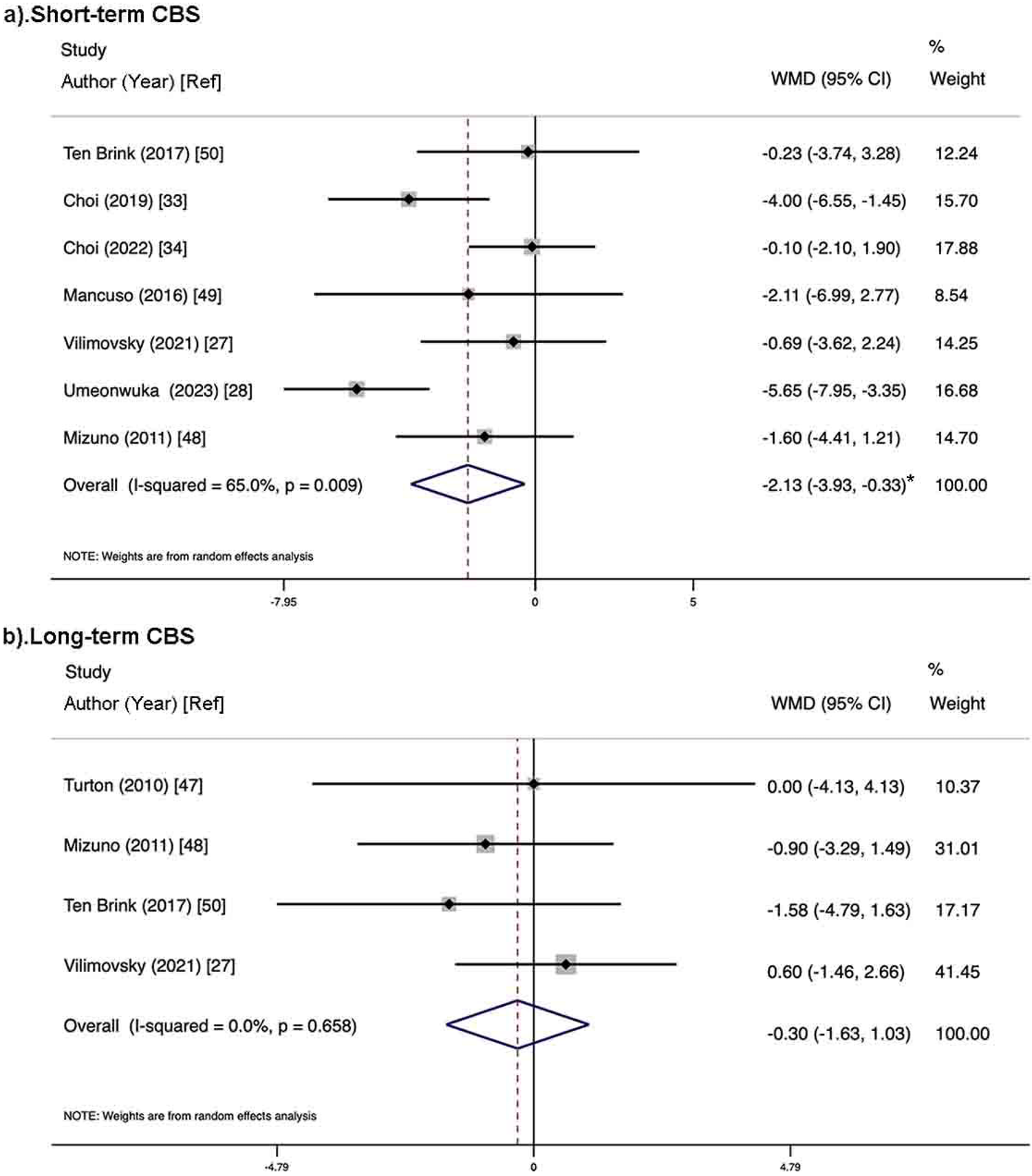
The effects of PA for main outcome-CBS. a). Short-term effects (7 studies included, n=288). The PA group showed significantly higher scores than controls (P <0.05), indicating PA’s immediate beneficial effect on CBS in SN participants. b). Long-term effects (four studies included, n=164), no significant long-term effects of PA on SN were observed (P <0.05). *: P <0.05; CBS: Catherine Bergego Scale; PA: prism adaptation; SN: spatial neglect.

For the long-term effect, four articles with a total of 164 participants were included, 80 in the PA group (average CBS score improvement: 6.68) and 84 in the control group (average CBS score improvement: 6.39)[27, 47, 48, 50]. The meta-analysis (Fig. 3b) found no significant difference between the two groups (WMD= −0.30, 95%CI: [−1.63, 1.03], P >0.05).

##### 3.3.1.1. Sensitivity analyses of the main outcome**—**CBS

According to the analyses in CBS, we observed high heterogeneity in the short-term analysis (I^2^ > 50%), while the heterogeneity of the long-term analysis was 0%. Influence analysis showed the research by Umeonwuka et al. (2023) was the main source of heterogeneity in short-term CBS. After exclusion, the heterogeneity (I^2^) of short-term CBS decreased from 65.0% to 21.2%, and the effects remained statistically significant (WMD = −1.38, 95%CI: [−2.70, −0.06], P <0.05). After further excluding the study by Choi et al. (2022), the heterogeneity (I^2^) decreased from 21.2% to 4%, with remaining significance (WMD = −1.92, 95%CI: [−3.34, −0.50], P <0.01), showing the robustness of this result. Therefore, we decided to retain those two studies (Umeonwuka et al. and Choi et al.) to maintain the integrity of the evidence.

#### 3.3.2 Secondary outcome—BIT

For the immediate effect for BIT-C, there were three articles with a total of 152 participants included, 78 in the PA group (average BIT-C score improvement: 25.40) and 74 in the control group (average BIT-C score improvement: 7.29)[28, 48, 49]. However, the meta-analysis didn’t reach statistical significance between the two groups (WMD= 17.84, 95% CI: [−4.87, 40.54], P >0.05; Fig. 4a). Only one study described BIT-B for immediate effects[48].

**Fig. 4.**
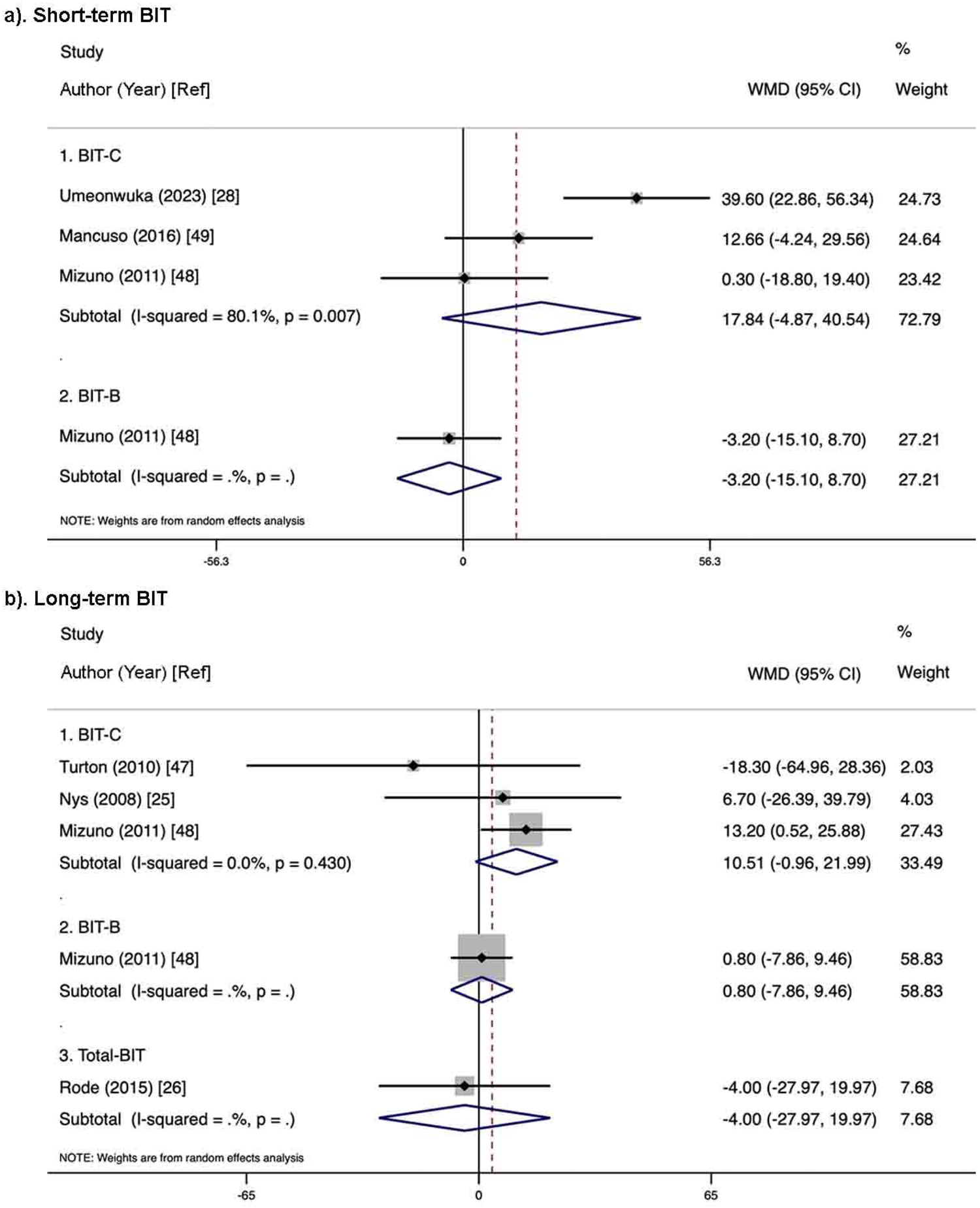
The effect of PA for secondary outcome-BIT. a). Short-term effects. For BIT-C (n=152, 3 studies), no significant difference was found between the PA group and the control group. Only one study reported BIT-B. b). Long-term effects. Three studies evaluated BIT-C (n=88), no significant long-term effects of PA on SN were observed. Limited data on BIT-B and total BIT from one study only. BIT: Behavioral Inattention Test; BIT-B: behavioral BIT; BIT-C: conventional BIT; PA: prism adaptation; SN: spatial neglect.

For the long-term effect, three articles evaluated BIT-C, including 88 participants in total—44 participants in both PA (average BIT-C score improvement: 46.83) and control (average BIT-C score improvement: 43.60) groups[25, 47, 48]. The meta-analysis found no significant difference between the two groups (WMD= 10.51, 95%CI: [−0.96, 21.99], P >0.05; Fig. 4b). Only one study each was included in BIT-B and Total BIT[26, 48].

##### 3.3.2.1 Sensitivity analyses of the secondary outcome**—**BIT

The study by Umeonwuka et al. (2023) explained most heterogeneity of the short-term CBS (I² decreased from 80.1% to 0% after its exclusion). However, the statistical conclusion of the retained two studies in short-term BIT-C remained non-significance (WMD= 7.23, 95% CI: [−5.42, 19.89], P >0.05). Therefore, we decided to retain this study.

For long-term BIT-C, although no statistical heterogeneity was observed (I² = 0%), influence analysis showed that the study by Mizuno et al. (2011) had little impact on combined results, as its exclusion maintained the effect non-significant (WMD = −1.67, 95%CI: [−28.66, 25.33], P >0.05). In addition, leave-one-out analysis found that excluding the study by Turton et al. (2010) altered the effect size to statistically significant (WMD = 12.37, 95% CI: [0.53, 24.21], P <0.05).

### 3.4 Univariate linear regression analyses for the main outcome—CBS

The regression analyses results revealed that there were no significant correlations between the age/days after stroke and PA intervention improvements (Table S2). None of the PA intervention parameters reached statistical significance either (Table S2). The scatter plot (Fig. S1) shows the improvements in the score of CBS benefits found for PA intervention as a function of physical intervention parameters.

### 3.5 Subgroup analyses for the main outcome—CBS

For the effect of prism shift, all seven studies included in short-term CBS utilized larger prism shift (≥10°), so results are identical to the findings in Section 3.3.1 (WMD = −2.13, 95%CI: [−3.93, −0.33], P <0.05)[27, 28, 33, 34, 48–50]. Only one study was included in the long-term CBS when the prism shift was <10°, while three studies had a prism shift of ≥10° and showed a non-significant difference (WMD = −0.34, 95%CI: [−1.74, 1.07], P >0.05; Fig. 5b)[27, 47, 48, 50].

**Fig. 5.**
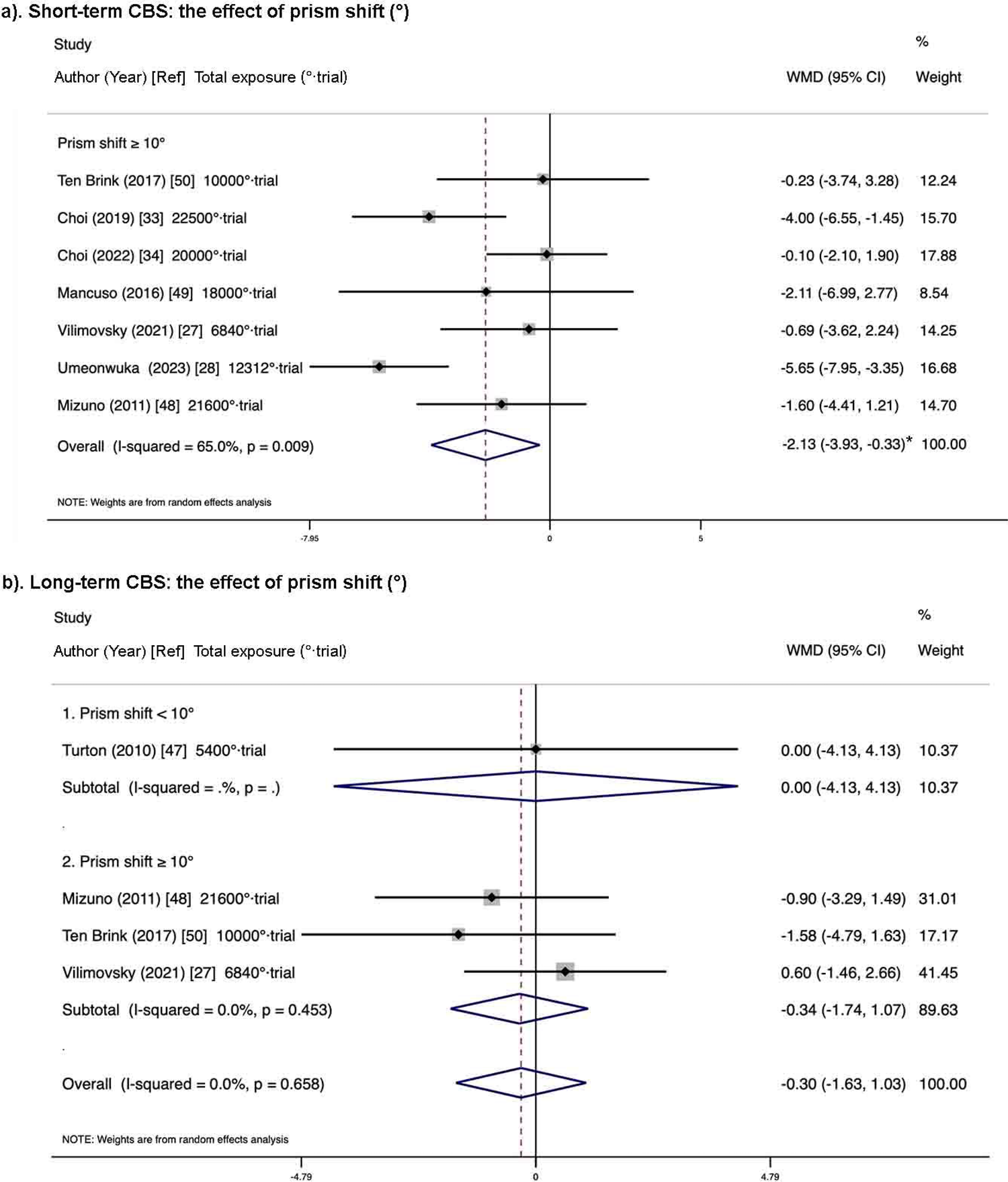
The subgroup analyses for the effects of prism shift (°) on CBS. a). Short-term subgroup analysis. All included seven studies (n=288, 7 studies) with ≥10° prism shift, and showed significant improvement between two groups (P >0.05). b). Long-term subgroup analysis. Only one study included in prism shift <10° subgroup, while other three studies (n=130) with ≥10° showed non-significance. *: P <0.05; CBS: Catherine Bergego Scale.

Based on the subgroup results for the total exposure, in short-term CBS, two studies were included in the subgroup of <11250°·trials and showed non-significance (WMD = −0.50, 95%CI: [−2.75, 1.75], P >0.05; Fig. 6a)[27, 50]. Five studies were included in the ≥11250°·trials subgroup and showed statistical significance (WMD = −2.73, 95%CI: [−5.01, −0.44], P <0.05; Fig. 6a)[28, 33, 34, 48, 49]. After excluding the studies by Umeonwuka et al. (2023) and Choi et al. (2022) to address heterogeneity, the effect size remained significant (WMD = −2.81, 95%CI: [−4.57, −1.05], P <0.01). For long-term CBS, there were three studies included in the <11250°·trials subgroup and showed non-significance (WMD = −0.03, 95%CI: [−1.63, 1.57], P >0.05; Fig. 6b)[27, 47, 50]; only one study was included in the ≥11250°·trials subgroup[48].

**Fig. 6.**
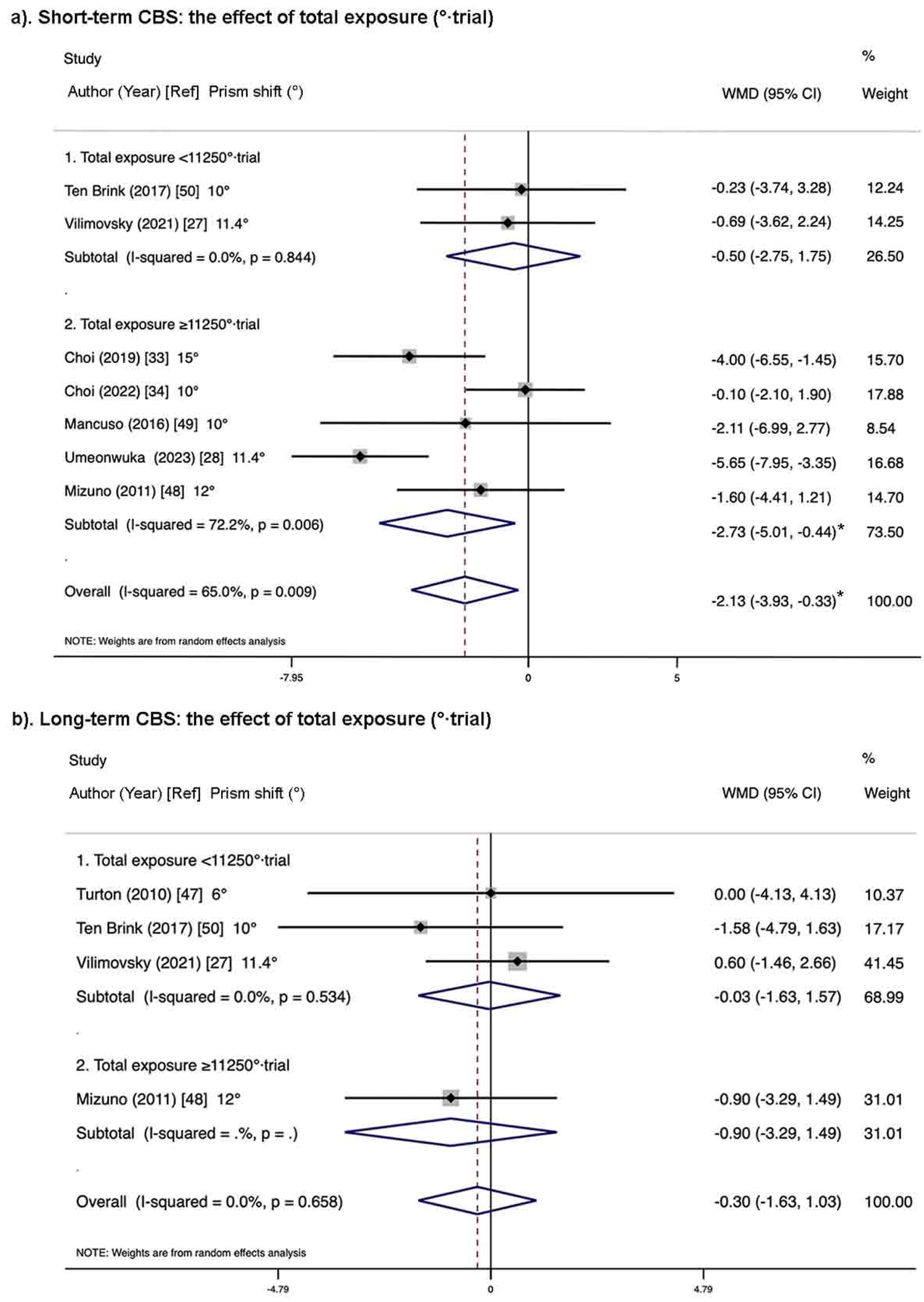
The subgroup analyses for the effects of total quantity of prism exposure (°·trial) on CBS. a). Short-term subgroup analysis. Two studies (n=92) included in <11250°·trial subgroup, showed non-significance; Three studies (n=196) with ≥11250°·trial showed significant improvement (P <0.05). b). Long-term subgroup analysis. Three studies (n=126) included in <11250°·trial subgroup, showed non-significance; Only one study with ≥11250°·trial. *: P <0.05; CBS: Catherine Bergego Scale.

## 4 Discussion

Our main aim was to provide a robust and reliable meta-analysis of RCTs investigating the effects of PA on SN. Compared with the four previous meta-analyses on the similar topic, we included the latest RCTs in this field up to March 2025, according to strict inclusion criteria. We distinguished between the short-term (i.e. <24 hours) and long-term (i.e. >1 month) effects of PA on CBS and BIT. Taken altogether, a significant immediate benefit was shown in CBS. The pooled effect on long-term BIT-C was influenced by the prism regime. No significant difference was observed for either the long-term CBS or the short-term BIT-C. For BIT-B, there was only one article providing data for the immediate efficacy and one for the long-term effect of BIT-B and total BIT, which was insufficient to assess effectiveness.

This study also discussed the potential predictive factor of PA efficacy that could account for result variability, and also preliminarily discussed the correlation of PA efficacy with participant characteristics (especially brain damage) in SN. To our knowledge, we provide the first meta-analysis that simultaneously analyzes the effect of assessment time, PA intervention parameters, and patient characteristics on PA intervention effects. Therefore, we hope that this study can provide preliminary exploratory insights into the “dose-response” mechanism of PA intervention and provide a reference for further research directions. The following discussion will first address the global meta-analysis results on CBS and BIT and then discuss predictive parameters for PA efficacy.

### 4.1 The effect of PA on SN

#### 4.1.1 Potential source of heterogeneity

In terms of the effect of PA on SN, analyses of both short-term CBS and short-term BIT showed relatively high heterogeneity (I^2^ > 50%). The study by Umeonwuka et al. was the main source of the heterogeneity in both short-term CBS and short-term BIT-C. The heterogeneity may stem from its characteristics that were different from those of other studies, e.g., a different design characteristic (single-center), younger participants (average age: 49.15 ± 13.31 years), fewer days after stroke (average: 39.25 ± 7.25 days), and the PA intervention parameters (Table 1). The study by Choi et al. also contributed to the heterogeneity of short-term CBS, presumably because most of the participants in this study were in the chronic stage of stroke (mean days after stroke: 279.30 ± 87.35 vs. <90 days in others). However, excluding these two studies had no impact on the conclusions. Based on the characteristics of the remaining studies, and combining the results of sensitivity and regression analyses, it was not feasible to fully identify a specific influencing factor for short-term efficacy. Interestingly, the results of the subgroup analysis provided some reference value (see the Physical predictive factors of PA efficacy section).

The effect size of long-term BIT-C was affected by Turton et al.’s study, possibly because of its smaller prism shift (6°). After its exclusion, the effect sizes of the remaining two articles (prism shift ≥10°) showed statistical significance. This trend suggested that the small prism shift used in this study may have weakened the overall effect of long-term BIT-C.

#### 4.1.2 short-term effect of PA on SN

The CBS and BIT scales were used to assess different aspects of the affected individuals with SN. For CBS, we obtained a significant short-term benefit of PA intervention, supporting the short-term efficacy of PA in ADL for participants with SN. The effects of PA intervention may thus translate into improvements in ADL for participants with SN[51].

For BIT-C, three studies were included in this meta-analysis[28, 48, 49]. In the study by Umeonwuka et al., they found that the BIT-C scores of the PA intervention group were significantly higher than those of the control group[28]. In the second study by Mizuno et al., they interestingly split the participants based on the SN severity and evaluated the effect of PA in BIT-C separately[48]. They found an obvious effect of PA on neglect symptoms and ADL in mild SN but no significant effect in severe SN. When analyzing the pooled group, the observed positive trend did not reach statistical significance. In another study following a similar procedure, they found that both groups showed a significant improvement in outcomes, but no statistical difference between the groups, even though the experimental group had higher improvement in all outcomes[49]. Overall, only three studies reported the immediate effect of PA on SN in terms of BIT-C, and the small sample size may increase the risk of undetected effects due to insufficient power. More large sample sizes and multicenter studies are needed to validate the results in the future.

#### 4.1.3 Long-term effect of PA on SN

In the global meta-analysis, no significant long-term effects of PA on SN were observed in CBS and BIT. This result is similar to the meta-analysis results of Li et al. and Qiu et al[29, 31]. The small sample size (a limitation also noted in the meta-analyses by Li et al. and Qiu et al.), may reduce statistical power[29, 31]. Although a significant improvement for BIT-C was observed when the prism shift was ≥10° (n=2), the sample size remained limited. We therefore agree with their conclusion that more rigorous studies with larger sample sizes exploring the dose of PA interventions are needed in the future to determine its benefits.

### 4.2 Other outcomes for the effect of PA on SN

Practically, each of the 10 RCTs included at least 2 outcomes assessing the PA efficacy. Except for the most frequently reported outcomes—CBS and BIT, there are also some other meaningful behavioral or neuropsychological outcomes for evaluating the PA efficacy reported in some of these 10 RCTs (such as FIM, Albert Test, Bells Test, Wheelchair Navigation Task, and scene copying task). These outcomes mainly focus on the evaluation of independence in daily life, visual-spatial attention, cognitive-motor coordination, and spatial perception for participants with SN. However, since the number of articles reporting these outcomes is insufficient for meta-analysis (n ≤ 2), we only focused on CBS and BIT.

In future RCT studies, more attention could be paid to diverse, highly reliable and sensitive outcomes to assess the treatment effect of PA on SN. For example, the Albert test and the Bells test are highly sensitive to SN and are able to detect the changes in participants’ attention allocation after PA intervention[52, 53]. Dynamic tasks and situational simulation-type tests—such as gardening or flea-market tasks, and Wheelchair Navigation—are also helpful in assessing spatial attention in a dynamic environment in people with SN[16, 54]. Virtual Reality (VR) Neglect Tests could simulate real-life scenarios through VR technology to measure people’s allocation of attention on the neglect side[55]. When sample size and expected effect sizes allow, evaluation across multiple different tests is more sensitive than any single test alone and should also be encouraged in future studies[53, 56].

### 4.3 Physical predictors of PA efficacy

Two main types of parameters may be involved in the PA interventions efficacy. First, the physical parameters and regime of prism exposure conditions affect sensorimotor after-effects and should therefore affect in turn the expansion of PA to therapeutic cognitive effects. Second, the individual deficit and lesion should also convey a predictive value of therapeutic interventions.

The sensorimotor effects of PA are closely related to the parameters of prism intervention, including the degree of prism shift, the number of trials/exposure duration (during prism exposure), frequency (/days), as well as the number of sessions and duration (days) of the PA intervention[10–12]. One should expect that the PA efficacy on SN also relates to some of these physical parameters[9, 57]. For example, Gammeri R et al. suggested that the visuomotor adaptation induced by low-power prisms (i.e., shifting the visual field by 5°, 6°, or 10°) might be insufficient to produce detectable effects in all participants. They argued that higher-power prisms (such as 10°, 12° or above) may be more likely to reach critical therapeutic thresholds, producing significant PA effects[9]. Similarly, Naito et al. (2025) suggested that a larger prism shift ≥10° was a crucial factor in eliciting prism adaptation effects[35]. However, in our analyses, we found that simply satisfying this parameter was not enough to guarantee efficacy. Specifically, for short-term CBS (all included articles with ≥10° prism shift), the effect size only made a significant benefit when the total exposure was ≥11250°·trials. We also observed that for the three included studies for short-term BIT-C, the prism shift was ≥10° and the total exposure was ≥11250°·trials. However, the results did not reach statistical significance maybe due to the limited sample size.

For the long-term effects of PA, the available data did not enable us to explore all combinations of outcomes, prism shift, and total exposure. For CBS, three studies were included in the prism shift ≥10° subgroup (two of which with <11250°·trials) and showed no statistically significant effect. For BIT-C, two studies with ≥10° showed significant improvement (identical to the finding after excluding Turton et al. in Section 3.3.2.1), which points to the important role of the larger prism shift. However, it was not available to determine the independent contribution of the larger total exposure due to its substantial disparity across those two studies (with 4000°·trials and 21600°·trials). For the effect of the total exposure, three and two studies with <11250°·trials were included for CBS and BIT-C, respectively, and both leading to non-significant results (the result for BIT-C was identical to the finding after excluding Mizuno et al. in Section 3.3.2.1). The effect of larger total exposure in long-term CBS and BIT-C was uncertain in our analysis because each included only one study. The existing evidence is insufficient to confirm the long-term clinical benefits of a larger total exposure. There is still a lack of data to explore the effects of smaller prism shifts. Our subgroup analysis of short-term CBS suggested that the variable of the total exposure warrants further exploration in future research.

The importance of cumulative dose has also been confirmed in other studies. Frassinetti et al. (not RCT) conducted a 2-week PA intervention on SN participants (with a 10° shift, 180 trials-20 mins/session, twice/day, 20 sessions in total, 36000°·trials for the total exposure), and the improvements in SN were still observed 5 weeks after the intervention[58]. But it is unclear whether this effect lasted beyond 5 weeks because the study did not include longer-term follow-up. In addition, Shiraishi et al. conducted an 8-week PA intervention (with 15° shifting, 50 mins/session, 4.2 times per week) in SN participants and observed that the effects lasted up to 6 weeks after prism removal[59]. Notably, in this latter study, instead of pointing performance, various activities were performed while exposed to the prism glasses: toss rings (10-15 min), pegboard exercise (10-15 min), and a ball throwing/dart game (20-30 min). Therefore, not only pointing performance but also various activities while wearing prismatic glasses may be important factors in observing the effects of PA on SN[59]. In summary, this study combined a stronger-than-average prism strength, a large number of sessions and greater and more ecological activities during prism exposure, leading to a long-lasting benefit. Finally, we may only preliminarily speculate that a larger total exposure and diversifying task arrangements should be encouraged in future studies to expect long-term beneficial effects in participants with SN.

In addition, the combination of different interventions may be more effective in treating SN than a single intervention[54, 60–62]. For example, in the studies of Hyun-Se Choi et al. in 2019 and 2022, they reported separately that the effect of PA intervention combined with functional electrical stimulation (FES)/neck vibration (NV) on SN was better than that of PA or FES/NV alone[33, 34]. However, they did not follow up to observe the long-term effects of PA on SN. Hence, in future studies, more high-quality, longer-term RCTs are needed to observe the effect of PA on SN. It can be a separate prism intervention, including not only pointing tasks but also various activities while exposed to the prism glasses. PA combined with other interventions should also be investigated to combine several mechanisms of action.

### 4.4 Individual predictive factors of PA efficacy

Some studies have suggested that younger participants have better SN functional recovery after PA intervention[28, 63]. Earlier PA intervention after stroke may also improve participants’ response to PA[63–65]. However, according to our regression results, no significant correlations were found between the age or days after stroke and the benefits of PA intervention on SN. It is worth noting that we could only conduct analyses for them based on group mean values. Subgroup analyses of age/days post-stroke were also not undertaken because subgroup classifications could only follow broad criteria (e.g., middle-aged vs. elderly; acute, subacute vs. chronic), and only one study could be included in the chronic stage. More information at the individual level should also be encouraged in future studies to explore the relevance of such information to the efficacy of PA intervention on SN.

It is important to consider that the participants’ deficits and lesions may also affect the treatment outcome. One of the possible reasons for the discrepancy between case reports and RCT and for the variability of RCT outcomes is that responders and non-responders have been described. Numerous articles have reported the link between the improvement of symptoms and the characteristics of brain damage in people with SN. For example, Umeonwuka et al. reported that the BIT-C was not only related to the treatment allocation but also related to the cognitive ability (for each unit increase in cognitive ability (Mini-Mental State Examination score), the recovery rate increased by 1.5 after controlling for confounders)[28]. In another study, Kamakura CK et al. found that the BIT-C was also related to the degree of white matter lesions[66]. Among the studies included in our meta-analyses, several articles (e.g.,[25, 47, 49]) did not report and/or discuss participants’ cognitive abilities and/or specifics of brain damage. It is therefore possible that confounding variables contributed to the lack of significance observed.

Additionally, the PA efficacy is also correlated with the severity of SN. As shown in the RCT by Mizuno et al., PA intervention significantly improved neglect symptoms and ADL in participants with mild SN, but not in severe ones. This may be due to greater potential for brain plasticity and learning abilities in participants with mild SN than in severe ones[48]. The severity of SN is related to the brain injury area and volume[67, 68]. More extensive brain damage is more likely to impact multiple brain circuits, thereby reducing compensatory capacity and affecting the therapeutic effect of PA[68]. One study found that SN severity is associated with a widespread decline in functional connectivity of attention, motor and auditory network nodes to the contralateral brain, and that these changes were observed mainly in right-hemisphere stroke participants[67, 69]. Another functional magnetic resonance imaging study revealed that SN severity is linked to the decreased connectivity of the right basal forebrain to the brainstem and basal ganglia, coupled with increased connectivity of the left frontal, temporal, and parietal arousal and attention network regions[67, 70]. PA may improve neglect symptoms by rebalancing intra- and inter-hemispheric network activity via acting on intact nodes of the damaged network, homologous nodes in the undamaged hemisphere, or both[71–73]. The white matter integrity between hemispheres may be a key condition for the PA efficacy on SN[66, 74]. For example, Lunven et al. found that PA may improve SN by tapping sensorimotor or prefrontal circuits more anterior to the corpus callosum[75]. Additionally, structures such as medial temporal lobe structures, the anterior cerebellar cortex, and obviously, the occipital lobe also play a significant role in the effect of PA on SN[76–78]. Given the important role of cerebello-parietal connections in PA (contributing to both recalibration and realignment), it is not surprising that a similar key role of these connections has been found for the therapeutic effects of PA[13, 79]. In future studies, for a better understanding of the effects of PA on SN and the mechanisms behind it, it is necessary to define the severity of SN, the associated deficits, the brain injury aera, as well as the structural and functional characteristics of the brain’s connectivity[48, 51, 67]. It is also important for future studies to consider the effect of interactions between participants’ characteristics and PA intervention parameters.

## 5. Limitation

Although meta-analyses also have intrinsic limitations, they provide useful information about therapeutic efficacy[80]. The stringent RCT inclusion criteria applied in our study had some counterpart risks in terms of limitations. First, some high-quality reports may have been excluded. Subsequently, the small sample size may lower statistical power. This is one of the possible reasons why we did not find significant results for long-term CBS and short-term BIT and why we could not determine which parameters play a significant independent predictive role in the therapeutic effect of PA. It was also not feasible to examine the interaction between potential factors affecting treatment outcomes due to the limited sample size.

## 6. Conclusion

Given the current debate on PA, this meta-analysis applied stringent inclusion criteria. Despite the fact that only 10 RCTs were included in our study and only two outcomes (CBS and BIT) could be evaluated, we found a significant short-term benefit of PA on SN for the functional scale CBS (all studies with prism shift ≥10°), with effects mainly derived from studies with a total exposure of ≥11250°·trials. A significant improvement for long-term BIT-C was observed when the prism shift was ≥10° (n=2, with 4000°·trials and 21600°·trials). Long-term effects on CBS and short-term effects on BIT-C did not reach statistical significance. These results provide evidence for the therapeutic potential of PA intervention, and suggest that the total quantity of PA exposure may be a useful predictive factor for efficacy.

## Supporting information

Supplemental material 1: PRISMA_2020_checklist

## Data Availability

All data produced in the present study are available upon reasonable request to the authors

## Acknowledgments

The authors wish to thank Sonia Alouche, Anne Cheylus, Alexandre Foncelle, Eric Koun and Frederic Volland for their excellent and agreeable technical assistance.

## Abbreviations

ADL: activities of daily living
BIT: Behavioral Inattention Test
BIT-B: behavioral BIT
BIT-C: conventional BIT
CBS: Catherine Bergego Scale
CI: Confidence Intervals
FIM: Functional Independence Measure
FES: functional electrical stimulation
MBI: Modified Barthel Index
NV: neck vibration
PA: prism adaptation
RCT: randomized controlled trial
SN: spatial neglect
VR: Virtual Reality
WMD: Weighted Mean Difference.

## Author’s contribution

YuanLiang Zhu designed the study and wrote the protocol. Author YuanLiang Zhu, Francois Quesque and Yves Rossetti managed the literature searches and analyses. Authors YuanLiang Zhu, Daisuke Nishida, Eric Chabanat and Yves Rossetti undertook the statistical analysis, and author YuanLiang Zhu wrote the first draft of the manuscript. Daisuke Nishida, Francois Quesque, Sophie Jacquin-Courtois, Jacques. Luaute, Gilles Rode and Yves Rossetti reviewed and edited the manuscript. Francois Quesque and Yves Rossetti supervised the research. All authors contributed to and approved the final manuscript.

## List of all Additional Materials

1. Fig. S1: Scatter plot of linear regression-physical intervention parameters. Normalized benefit scores are pitted against two potential parameters. Each point corresponds to either short-term or long-term effects of PA on CBS benefit scores. The size of the points reflects the sample size of each study, the larger the sample size, the larger the point size (four bins were used: 10-19; 20-39; 40-59; 60-79). a). Prism strength: 9/11 data points exhibit better scores after PA intervention compared to the control group. b). Total quantity of prism exposure: 8/10 data points exhibit better scores). Although the regression line between outcome and prism shift/total quantity of prism exposure has a positive slope, as one may expect, this trend did not reach significance. CBS: Catherine Bergego Scale; PA: prism adaptation.
2. Table S1: The number of data points available for outcomes.
3. Table S2: The linear regression analyses-the predictive ability of participants’ basic characteristics (average age and days after stroke) and different PA intervention parameters on SN improvement (main outcome-CBS).
4. PRISMA checklist.

