## Supplemental Table 1 for "Robust immediate and limited long-term benefit of prism adaptation on spatial neglect: A Systematic Review and Meta-Analysis of outcomes and predictors"

Table S1. The details of number of reports for each outcome indicator

| **Outcomes type** | **Outcomes indicator** | **Short-term**  **(number of reports)** | **Long-term**  **(number of reports)** |
| --- | --- | --- | --- |
| Main outcome-CBS | CBS | 7 | 4 |
| Secondary outcome-BIT | BIT-C | 3 | 3 |
|  | BIT-B | 1 | 1 |
|  | Total-BIT | NA | 1 |
| Other outcomes | Albert Test | 2 | NA |
|  | Bells Test | 2 | 1 |
|  | Letter Cancellation | 2 | NA |
|  | Line Bisection | 2 | NA |
|  | FIM | 1 | 2 |
|  | MBI | 1 | 1 |
|  | Wheelchair Navigation Task | 1 | 1 |
|  | Others | ≤1 | ≤1 |

BIT: Behavioral Inattention Test; BIT-B: behavioral BIT; BIT-C: conventional BIT; CBS: Catherine Bergego Scale; FIM: Functional Independence Measure; MBI: Modified Barthel Index.
