## Supplemental Table 2 for "Robust immediate and limited long-term benefit of prism adaptation on spatial neglect: A Systematic Review and Meta-Analysis of outcomes and predictors"

|  | **Parameters** | **Slope** | **Intercept** | **R** | **t** | **P** |
| --- | --- | --- | --- | --- | --- | --- |
| **Participates characteristics** | Age (year) | -0.890 | 0.641 | 0.420 | -1.390 | 0.198 |
|  | Days after stroke | 0.014 | 0.089 | 0.053 | 0.160 | 0.876 |
| **PA intervention parameters** | Prism shift (°) | 2.987 | -24.282 | 0.374 | 1.210 | 0.257 |
|  | Number of trials | -0.228 | 28.011 | 0.183 | -0.525 | 0.614 |
|  | Number of sessions | 0.923 | -5.131 | 0.289 | 0.906 | 0.389 |
|  | Number of the total trials* | 0.008 | -2.711 | 0.233 | 0.678 | 0.517 |
|  | Total quantity of prism exposure^#^ | 8.276 x 10^-4^ | -3.491 | 0.320 | 0.957 | 0.367 |
|  | Frequency (/days) | 2.677 | 4.120 | 0.083 | 0.250 | 0.808 |
|  | Number of PA intervention days | 2.174 | -16.625 | 0.375 | 1.215 | 0.255 |
|  | Total duration of PA intervention (days elapsed between the first-to-last sessions) | 1.861 | -18.044 | 0.463 | 1.568 | 0.151 |

Table S2. The linear regression analysis- the predictive ability of different PA intervention parameters on SN improvement (main outcome-CBS).

*: Number of the total trials = (number of the trials x number of sessions)

^#^: Total quantity of prism exposure = (number of total trials x prism shift)

BIT-C: conventional BIT; CBS: Catherine Bergego Scale; PA: prism adaptation; SN: spatial neglect.
