## Supplementary figures and images for "Robust immediate and limited long-term benefit of prism adaptation on spatial neglect: A Systematic Review and Meta-Analysis of outcomes and predictors"

### Supplemental Figure 1

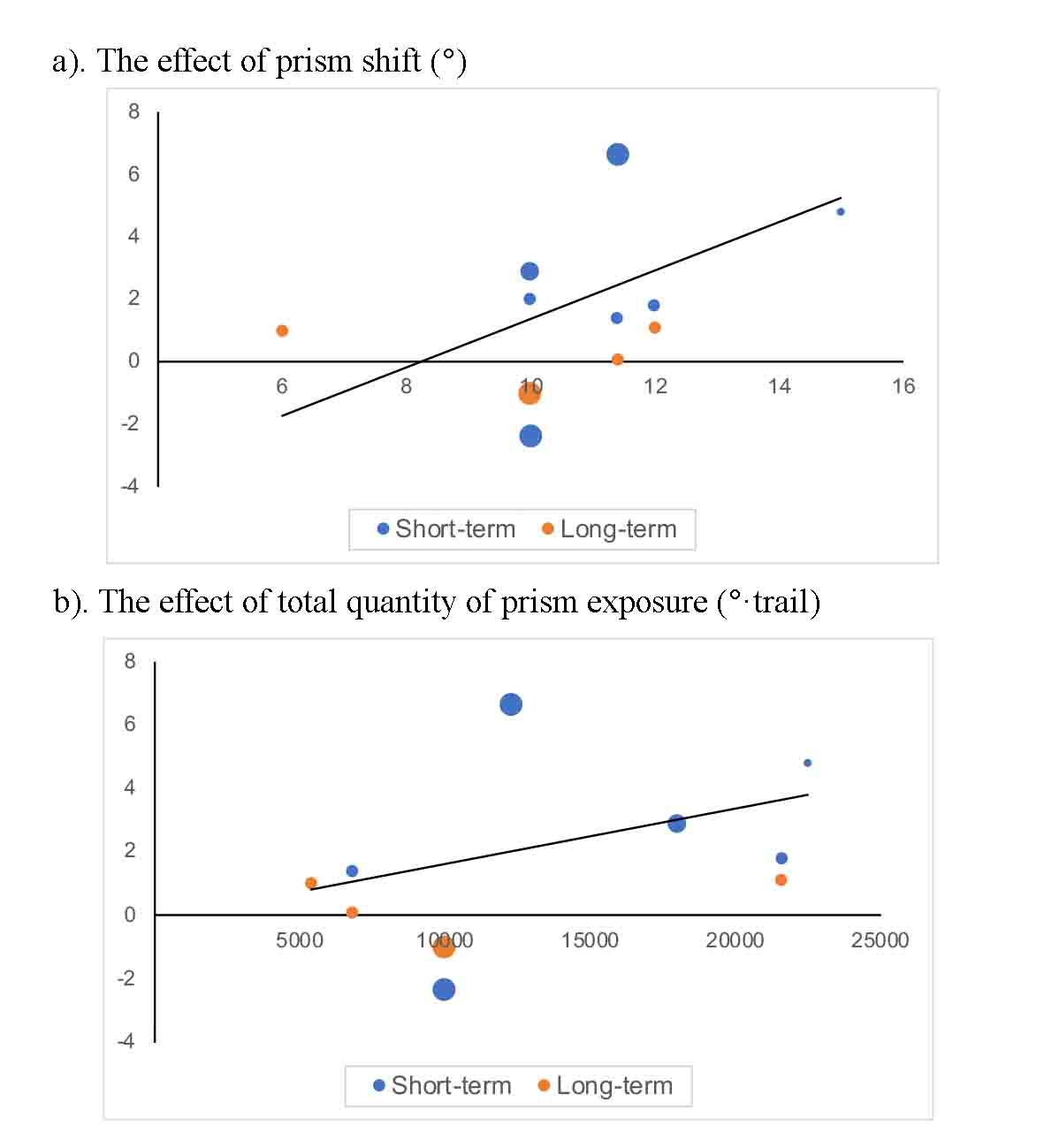
